# Optimizing Hypertension Management in Primary Aldosteronism Screening: The Role of Dual Calcium Channel Blockers

**DOI:** 10.1101/2025.10.13.25337950

**Authors:** Xingyu Jiang, WenChao Zhang, LiXia Wei, Yan Tang, Jiawen Pang, Shanshan Chen, Jianling Li

**Affiliations:** Department of Cardiovascular Medicine, The First Affiliated Hospital of Guangxi Medical University, Nanning, Guangxi 530021, China; Guangxi Medical University, Nanning, Guangxi 530021, China; Department of Cardiovascular Medicine, The Second People’s Hospital of Qinzhou, Qinzhou, Guangxi 535000, China

**Keywords:** Primary aldosteronism, Verapamil, Amlodipine, Dual Calcium Channel Blockers

## Abstract

Primary aldosteronism (PA) poses a distinct challenge in hypertension management, especially during the drug washout phase before diagnostic screening. This retrospective study evaluated the effectiveness and safety of a dual calcium channel blocker (CCB) regimen—amlodipine combined with verapamil—for blood pressure control during this critical period.

We reviewed data from 434 hypertensive patients who underwent secondary hypertension screening at the First Affiliated Hospital of Guangxi Medical University between 2019 and 2025. Compared with the antihypertensive treatments used before the washout phase, the dual CCB approach achieved significant reductions in seated systolic blood pressure (SBP) (163.08 ± 22.28 mmHg vs. 135.16 ± 14.85 mmHg; p < 0.001), diastolic blood pressure (DBP) (99.90 ± 15.96 mmHg vs. 83.38 ± 12.27 mmHg; p < 0.001), and heart rate (HR) (85.29 ± 15.61 bpm vs. 76.14 ± 9.42 bpm; p < 0.001). When comparing patients with PA to those with essential hypertension, there were no significant differences in SBP (132.51±14.16 vs. 135.41±14.57 mmHg; p = 0.276), DBP (81.24±12.12 vs. 82.47±10.94 mmHg; p = 0.562), or HR (75.71±7.61 vs. 76.34±11.17 bpm; p = 0.722). The only adverse event noted was intermittent first-degree atrioventricular block in one patient.

In conclusion, the dual CCB regimen proved both effective and well tolerated during the drug washout phase prior to PA screening. These findings highlight a potential strategy for optimizing blood pressure management in this context. Further studies are warranted to confirm its long-term role in clinical practice.

## Introduction

Recent research has shown that primary aldosteronism (PA) often develops gradually, beginning with a preclinical or subclinical stage before progressing to overt disease[1]. Excess aldosterone affects both the cardiovascular and renal systems, leading to combined damage over time[2]. Despite these risks, many patients remain symptom-free for years. Early detection is therefore critical: simplifying PA diagnosis and subtyping allows timely treatment, which can lower cardiovascular and renal complications in hypertensive patients. However, many eligible patients are never screened[3, 4], largely because of limited awareness, restricted access to resources, and shortcomings in current screening methods.

Current guidelines recommend the aldosterone-to-renin ratio (ARR) as the first-line screening test for PA. Yet ARR values can be altered by commonly prescribed antihypertensive drugs. Stopping these medications to obtain accurate screening results increases patient risk. To address this, drugs with minimal influence on ARR—such as non-dihydropyridine calcium channel blockers (non-DHP-CCBs) and alpha-blockers—are typically recommended during the washout phase. More recently, studies suggest that dihydropyridine calcium channel blockers (DHP-CCBs) also have little effect on ARR[5], offering another potential option. This highlights the need to further examine how antihypertensive therapy can be optimized during the washout period to balance patient safety with diagnostic accuracy.

Against this background, we performed a retrospective analysis of patients who underwent secondary hypertension screening at a single center between 2019 and 2025. The purpose of this study was to compare the effectiveness and safety of different antihypertensive regimens in managing blood pressure during the washout phase.

## Methods

### Data Collection

This study adhered to the ethical principles outlined in the Declaration of Helsinki and was approved by the Ethics Committee of the First Affiliated Hospital of Guangxi Medical University. As a retrospective study, the ethics committee waived the requirement for patient informed consent. The study population consisted of patients who underwent hypertension screening at the First Affiliated Hospital of Guangxi Medical University between 2019 and 2025. Exclusion criteria included patients who were unsuitable for medication washout procedures, such as those with severe infections, severe hepatic insufficiency, or those who were pregnant or lactating.

Data were extracted from the electronic medical records system of the First Affiliated Hospital of Guangxi Medical University. The information collected included demographic details such as patient age, sex, race, height, and weight. Additionally, clinical history data were gathered, including any history of hypertension, diabetes, hypokalemia, and adverse reactions following medication adjustments. Laboratory data were also collected, including lipid profiles, electrolyte levels, aldosterone-to-renin ratio (ARR), plasma aldosterone concentration (PAC), plasma renin activity (PRA), and other relevant laboratory markers.

Blood pressure and heart rate data were recorded both before and after medication adjustments. Detailed information on antihypertensive treatment regimens was also obtained, including the types, dosages, and treatment duration before and after medication adjustments. All collected data were anonymized and handled in compliance with data protection regulations to ensure patient privacy.

### Efficacy and Safety Evaluation

The primary efficacy assessment in this study focused on blood pressure control. Blood pressure was measured using a standardized, calibrated cuff-type sphygmomanometer to ensure accuracy and consistency. To minimize physiological fluctuations due to time factors and ensure data reliability and comparability, all blood pressure measurements were conducted at the same time each day. To further reduce the impact of random factors, measurements began on the second day after medication adjustment, with continuous blood pressure and heart rate monitoring performed daily. The average of these continuous measurements was then used as the final data, which effectively minimized errors from single measurements and provided more robust data on blood pressure fluctuations. Furthermore, all measurements were carried out by specially trained research personnel to ensure consistency and standardization in the procedures. The safety evaluation aimed to monitor and document all adverse events (AEs) and serious adverse events (SAEs) associated with medication adjustments. These adverse reactions included mild side effects, such as dizziness, fatigue, and gastrointestinal discomfort, as well as more serious reactions, including hypotension, arrhythmias, and bilateral lower limb edema.

### Statistical Methods

All data analyses were performed using R software version 4.2.2. Statistical tests were conducted with a two-sided hypothesis and a significance level set at P < 0.05. For continuous data that followed a normal distribution, normality was assessed using the Shapiro-Wilk test, and the data were expressed as mean ± standard deviation (Mean ± SD). For non-normally distributed continuous data, the data were presented as median and interquartile range (M [Q1, Q3]), with comparisons made using the Mann-Whitney U test. Categorical data were presented as frequency and percentage [n (%)] values. To minimize the potential influence of baseline characteristics, such as blood pressure, weight, and medication use, on the effects of antihypertensive treatment, propensity score matching (PSM) was applied. After matching, the balance between groups was assessed using the standardized mean difference (SMD).

## Results

This study included 434 patients with hypertension, all of whom underwent secondary hypertension screening. Clinicians selected the appropriate washout regimen according to each patient’s admission blood pressure and their clinical judgment. Medications prescribed during this period included amlodipine (5–10 mg daily), verapamil (120–240 mg daily), prazosin (5 mg daily), and terazosin (1 mg daily). The baseline characteristics of all patients are summarized in Table 1.

**Table 1.**
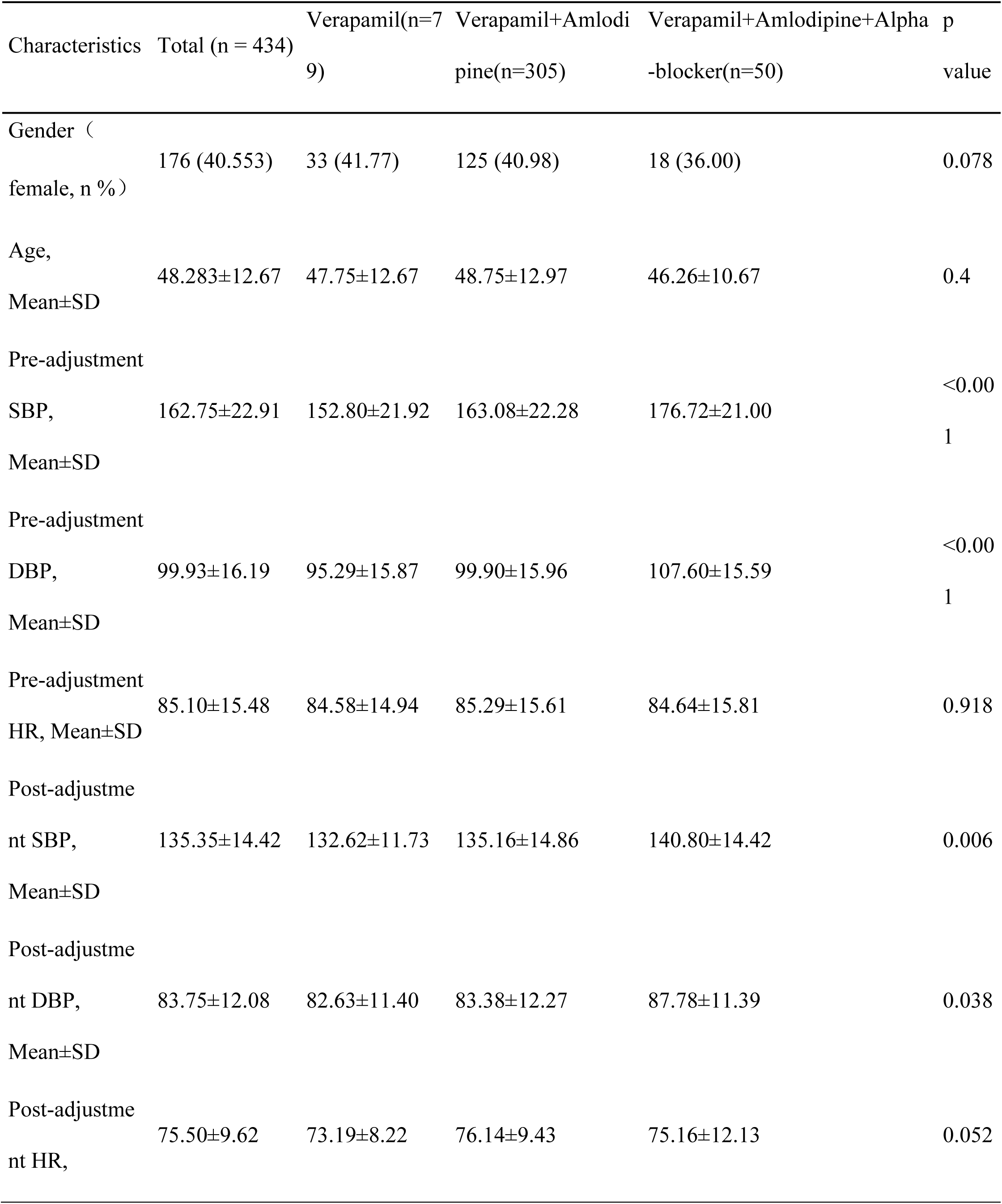
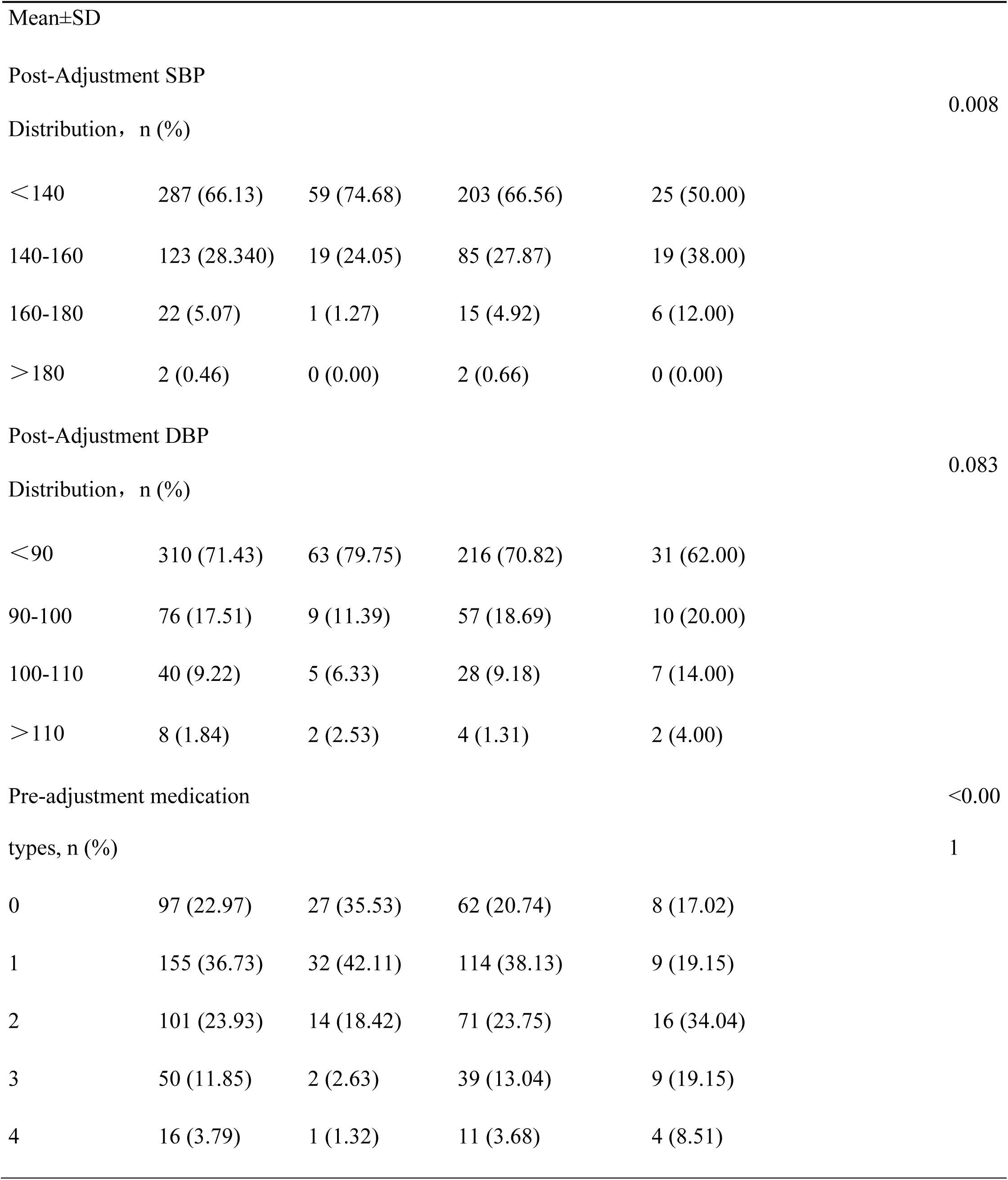
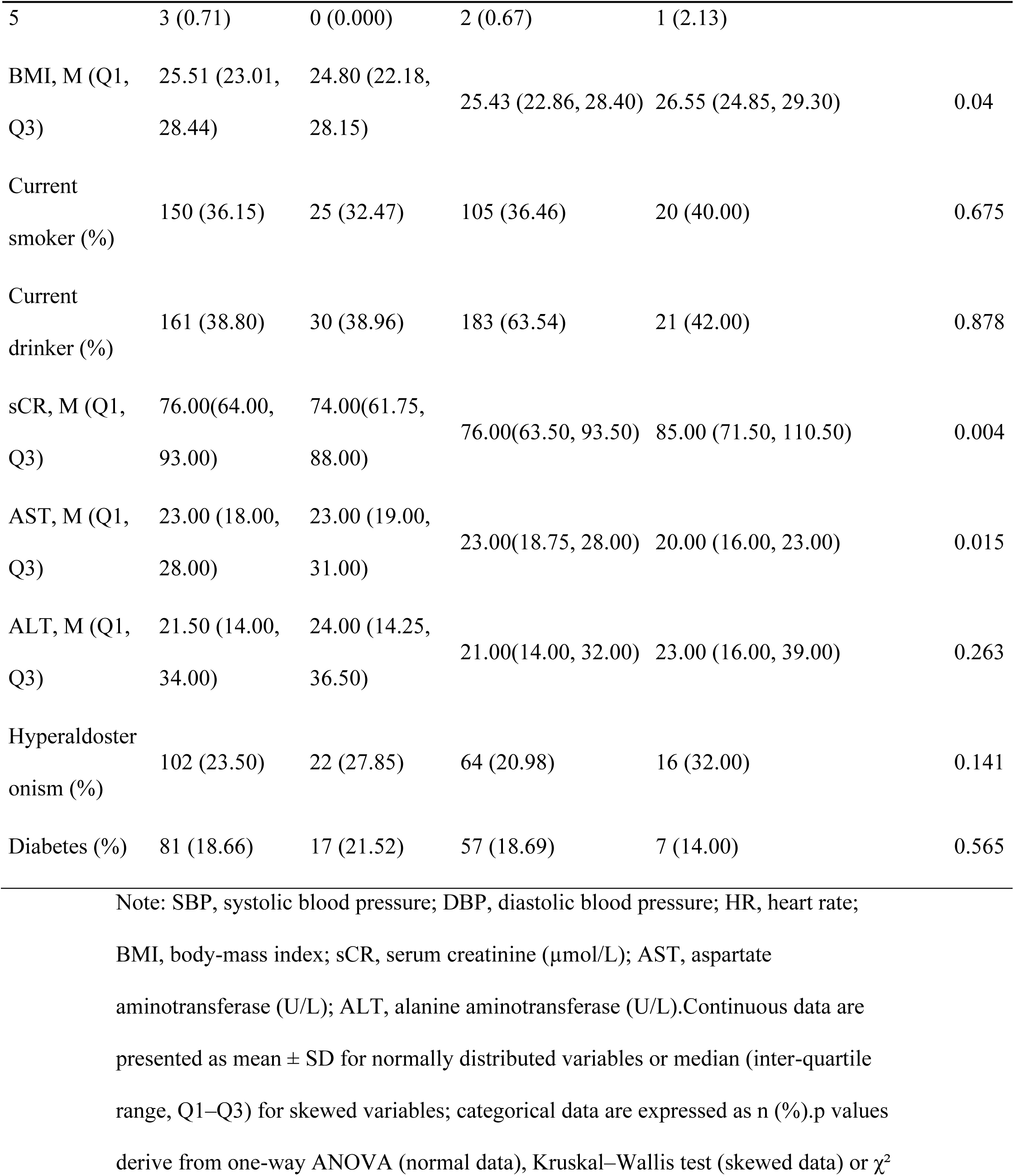
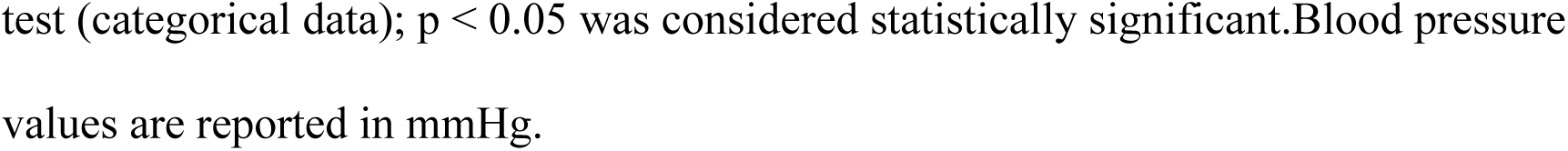
Baseline characteristics of study participants.

At baseline, patients receiving the three different regimens showed significant differences in blood pressure. Both the dual calcium channel blocker (CCB) group and the dual CCB plus alpha-blocker group had higher pre-adjustment systolic blood pressure (SBP) and diastolic blood pressure (DBP) compared with the verapamil group (p < 0.001). The highest baseline SBP was observed in the dual CCB plus alpha-blocker group (176.72 ± 21.00 mmHg), followed by the dual CCB group (163.08 ± 22.28 mmHg), while the verapamil group had the lowest mean SBP (152.80 ± 21.92 mmHg). After medication adjustment, no significant differences were observed between the dual CCB and verapamil groups in SBP (135.16 ± 14.85 vs. 132.62 ± 11.73 mmHg; p = 0.371), DBP (83.38 ± 12.27 vs. 82.63 ± 11.40 mmHg; p = 0.886), or heart rate (HR) (76.14 ± 9.42 vs. 73.19 ± 8.22 bpm; p = 0.052). Overall, the dual CCB group showed a stronger antihypertensive effect compared with verapamil alone.

To further explore the role of dual CCB therapy in PA screening, we used body mass index (BMI), pre-adjustment blood pressure, and medication type as covariates and conducted 1:1 propensity score matching. A total of 241 patients with essential hypertension (EH) and 64 patients with PA were matched using a caliper width of 0.1 standard deviation (SD). After matching, 59 patients with PA and 59 patients with EH were analyzed (Table 2). Post-adjustment outcomes showed no significant differences between the two groups in SBP (132.51±14.16 vs. 135.41±14.57 mmHg; p = 0.276), DBP (81.24±12.12 vs. 82.47±10.94 mmHg; p = 0.562), or HR (75.71±7.61 vs. 76.34±11.17 bpm; p = 0.722), suggesting that dual CCB therapy is equally effective for both PA and EH patients.

**Table 2.**
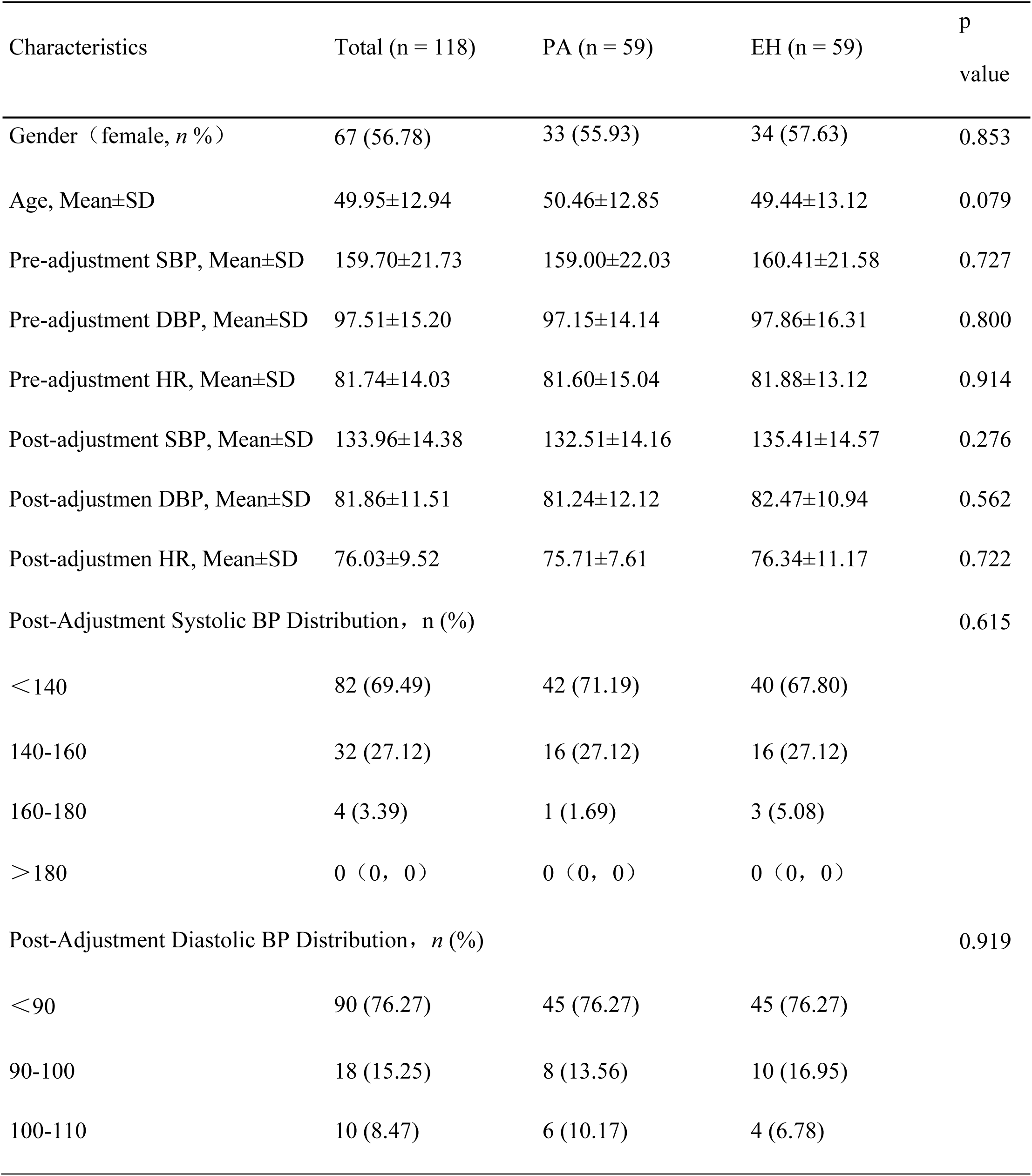
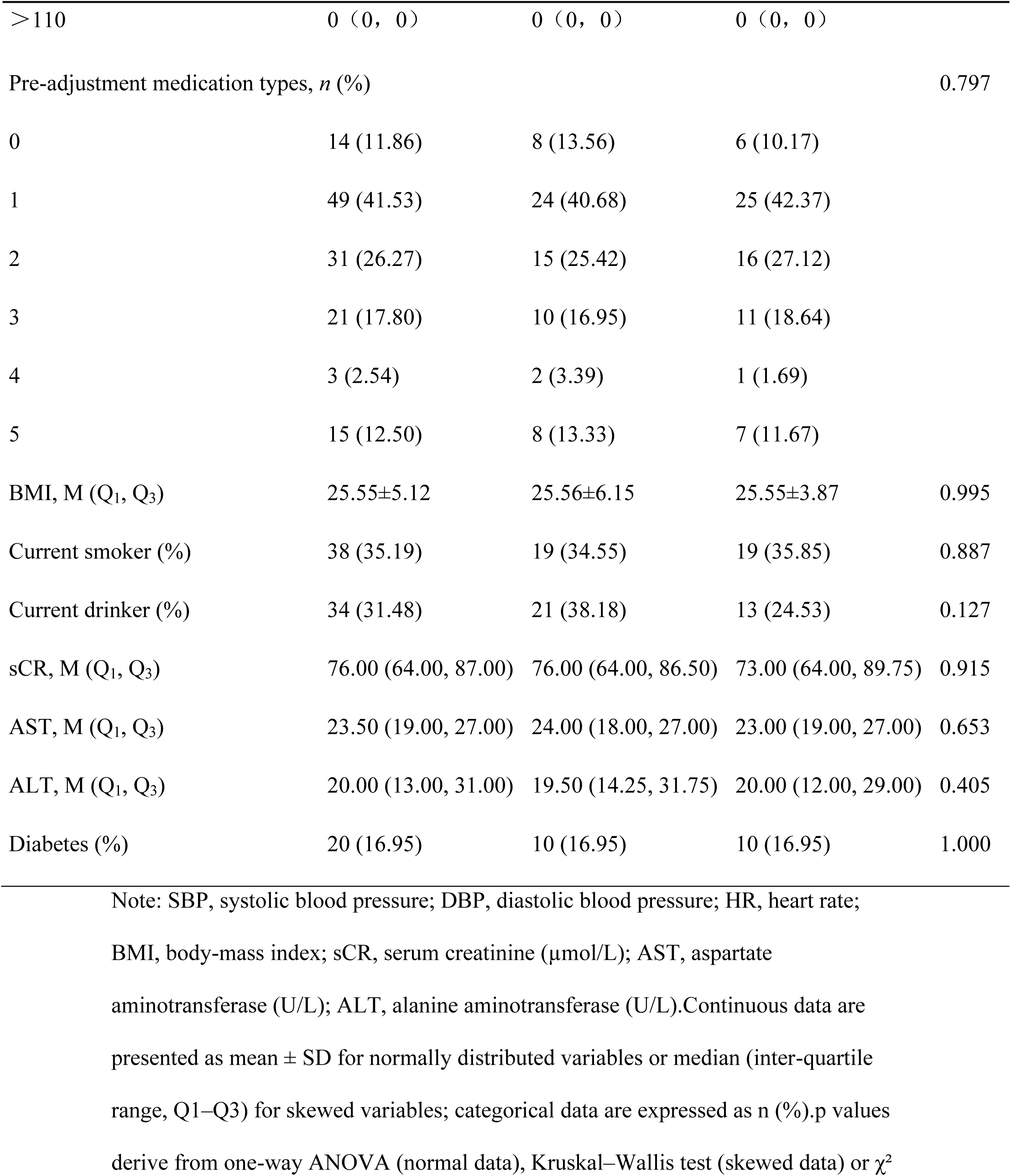
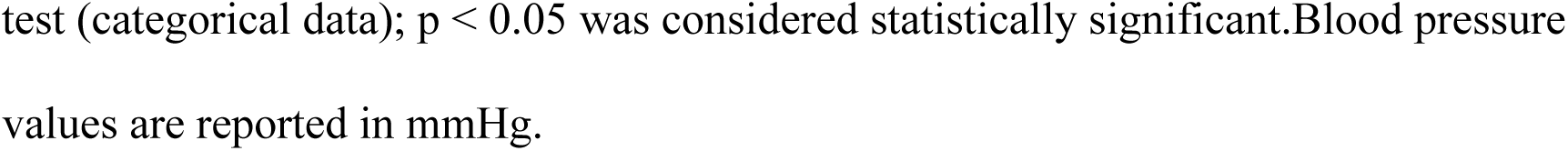
Dual CCB therapy in primary aldosteronism vs essential hypertension.

To compare dual CCB therapy with dual CCB plus alpha-blocker therapy, we again performed propensity score matching with BMI, pre-adjustment blood pressure, and medication type as covariates. Matching was conducted between 305 patients on dual CCB therapy and 50 patients receiving dual CCB with an alpha-blocker, with a caliper width of 0.1 SD. After matching, 42 patients in each group were analyzed. As shown in Table 3, no significant differences were found between the groups after adjustment in SBP (141.38 ± 13.44 vs. 140.24 ± 14.34 mmHg; p = 0.707), DBP (86.31 ± 10.24 vs. 87.14 ± 11.38 mmHg; p = 0.725), HR (75.93 ± 8.89 vs. 74.79 ± 12.85 bpm; p = 0.637), or in SBP and DBP distribution (p = 0.971 and p = 0.791, respectively). These results indicate that dual CCB therapy alone achieves comparable blood pressure and heart rate control to dual CCB combined with an alpha-blocker.

**Table 3.**
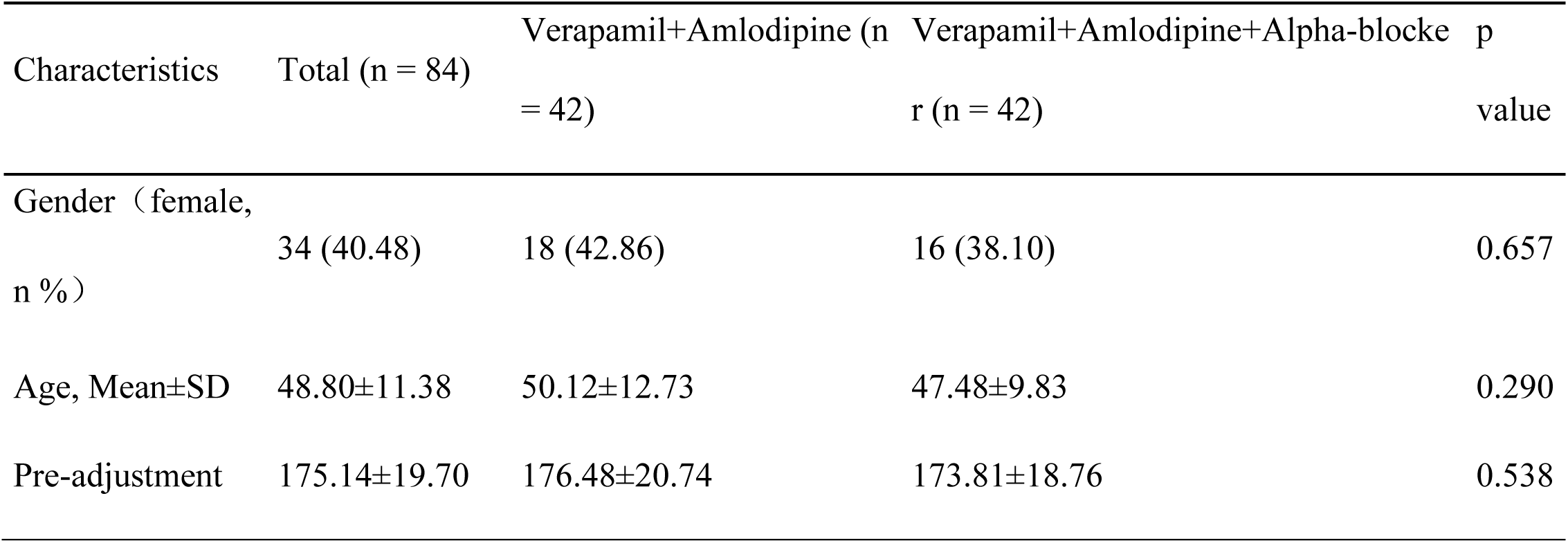
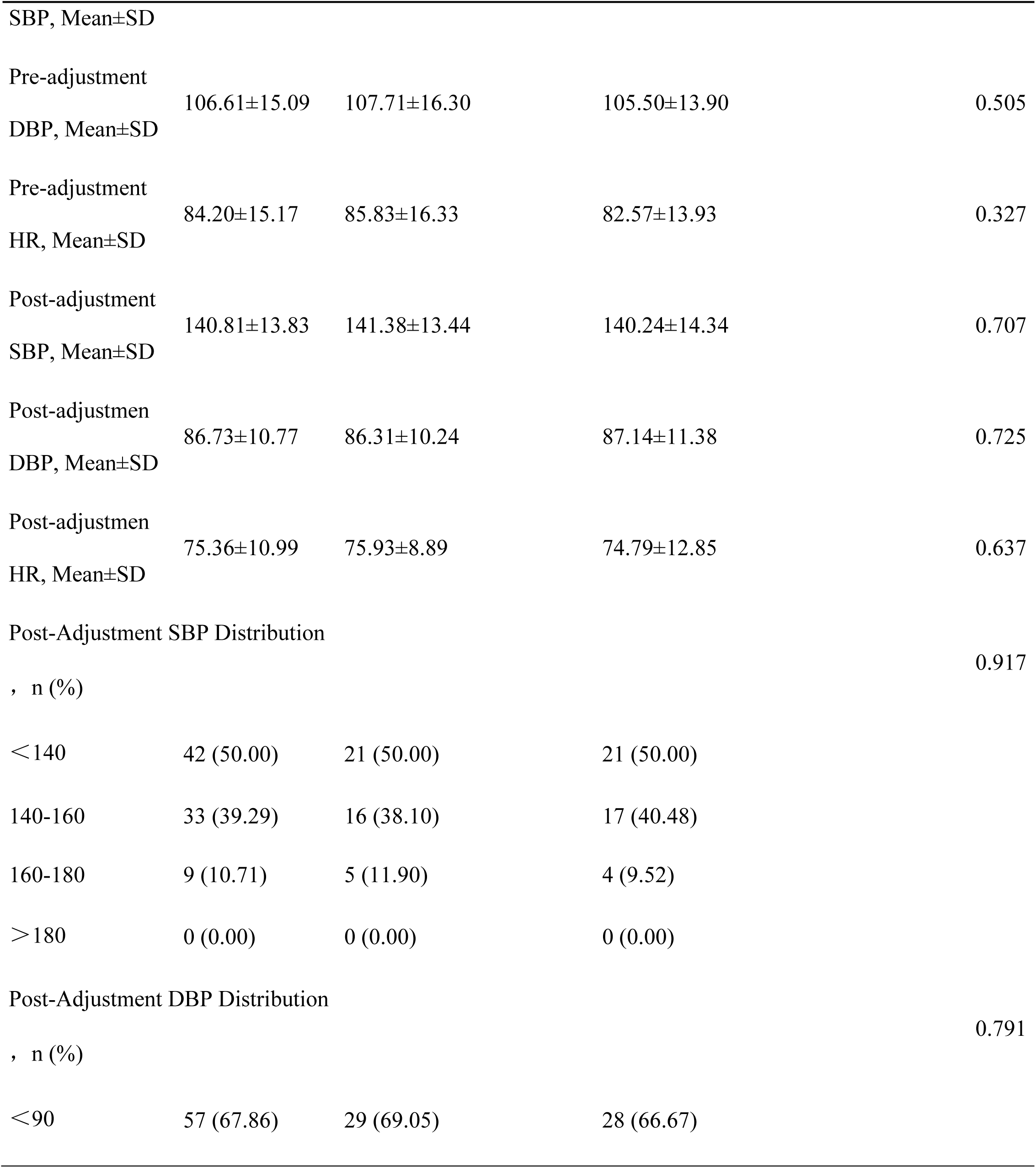
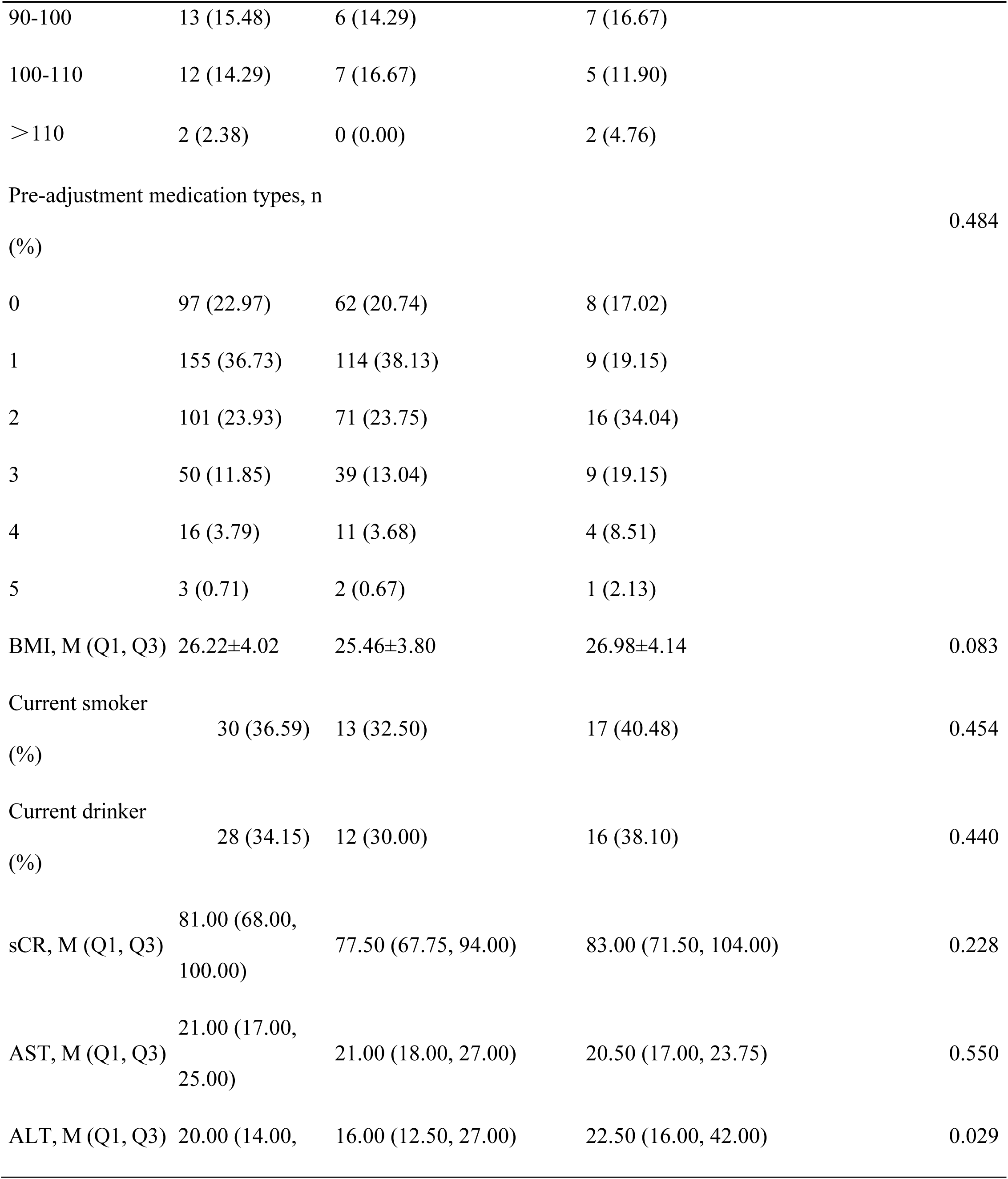
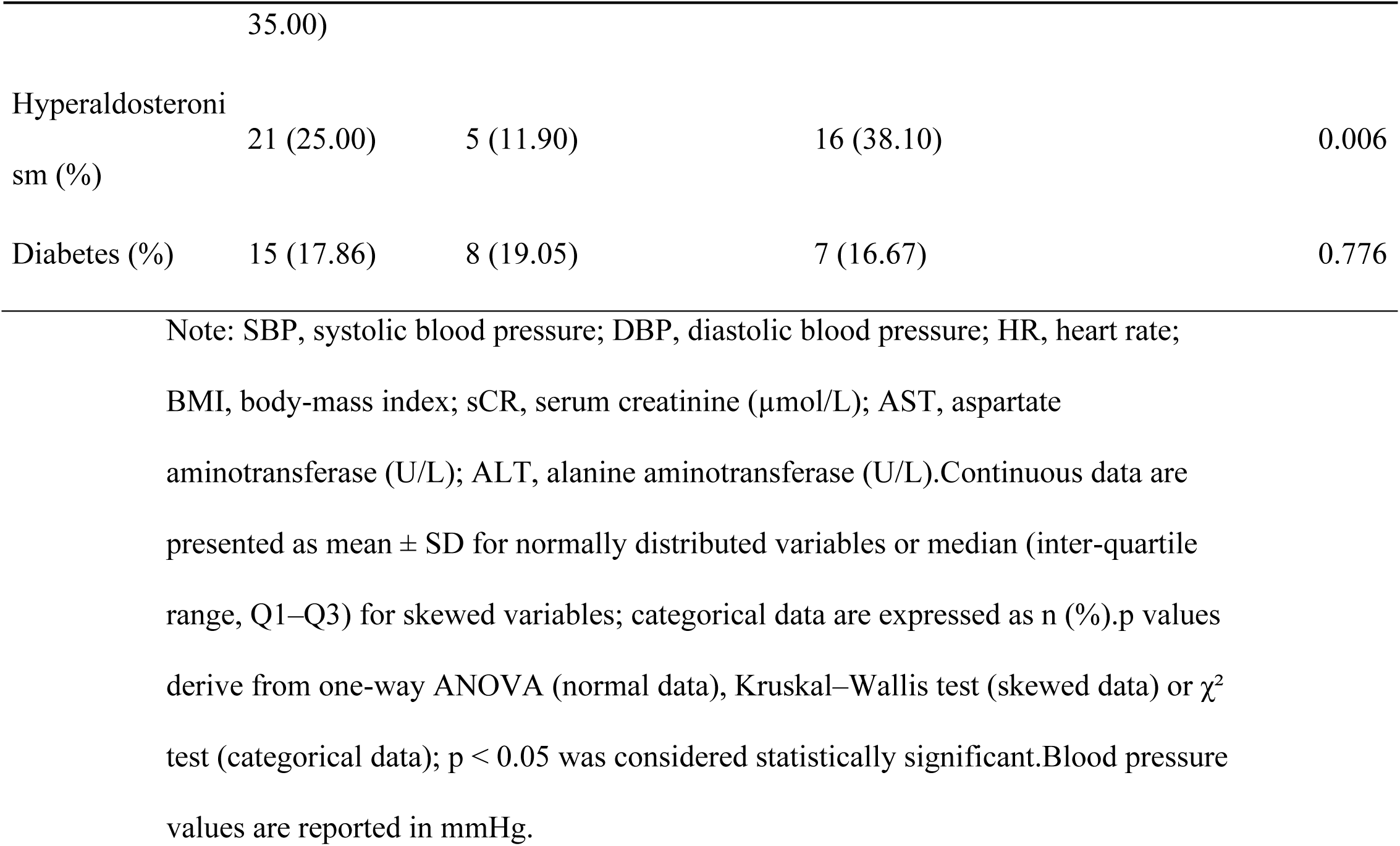
Comparison between dual CCB therapy and dual CCB therapy plus alpha-blocker.

A self-controlled analysis was performed to evaluate the effect of dual CCB therapy more directly (Table 4; Figure 1). After adjustment, patients on dual CCB therapy showed significant reductions in SBP (163.08 ± 22.28 vs. 135.16 ± 14.85 mmHg; p < 0.001), DBP (99.90 ± 15.96 vs. 83.38 ± 12.27 mmHg; p < 0.001), and HR (85.29 ± 15.61 vs. 76.14 ± 9.42 bpm; p < 0.001). Improvements were also observed in the distribution of both SBP and DBP (p < 0.001 for both).

**Figure 1.**
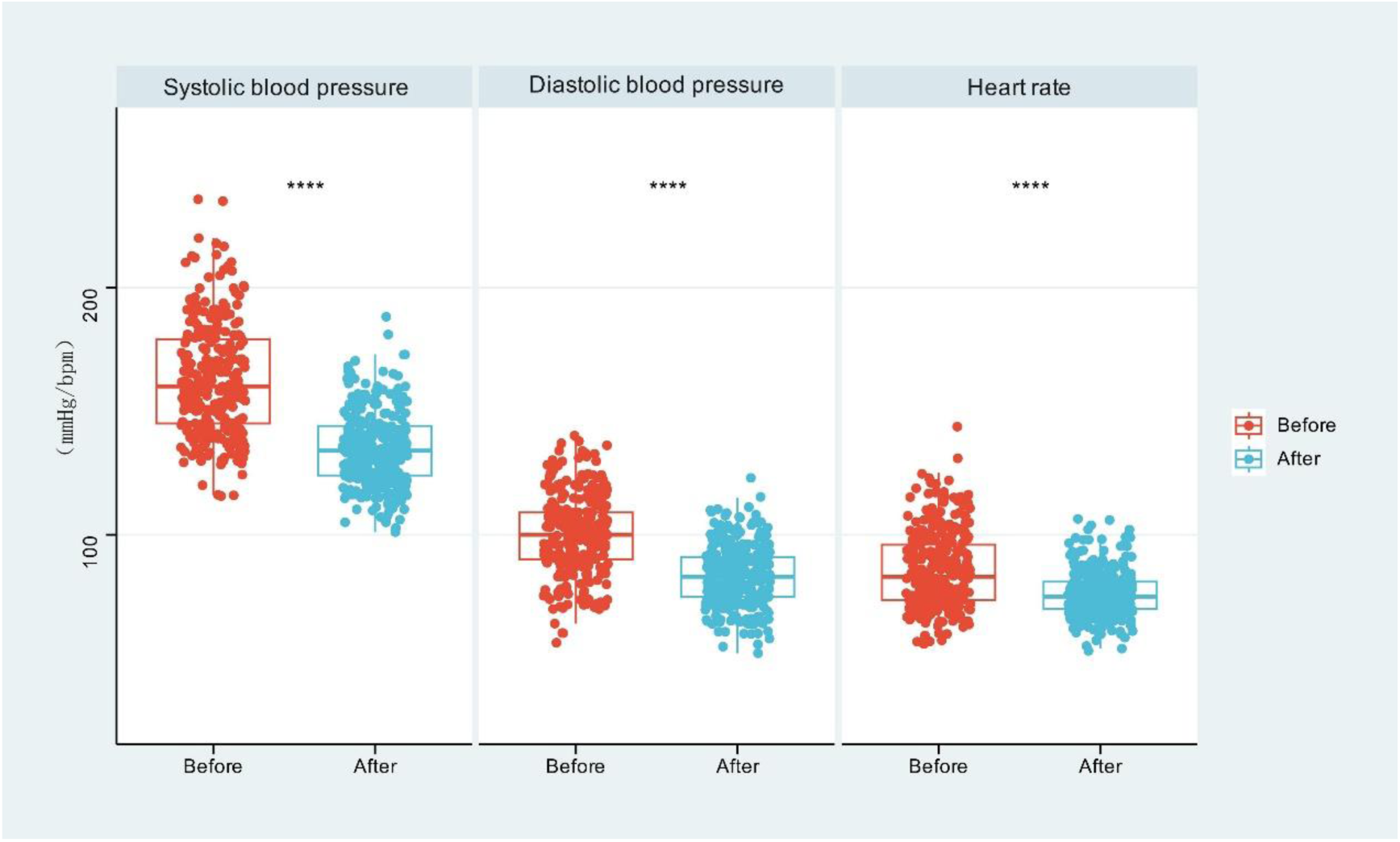
Blood Pressure Comparison Before and After Transition to Dual CCB Therapy.

**Table 4.**
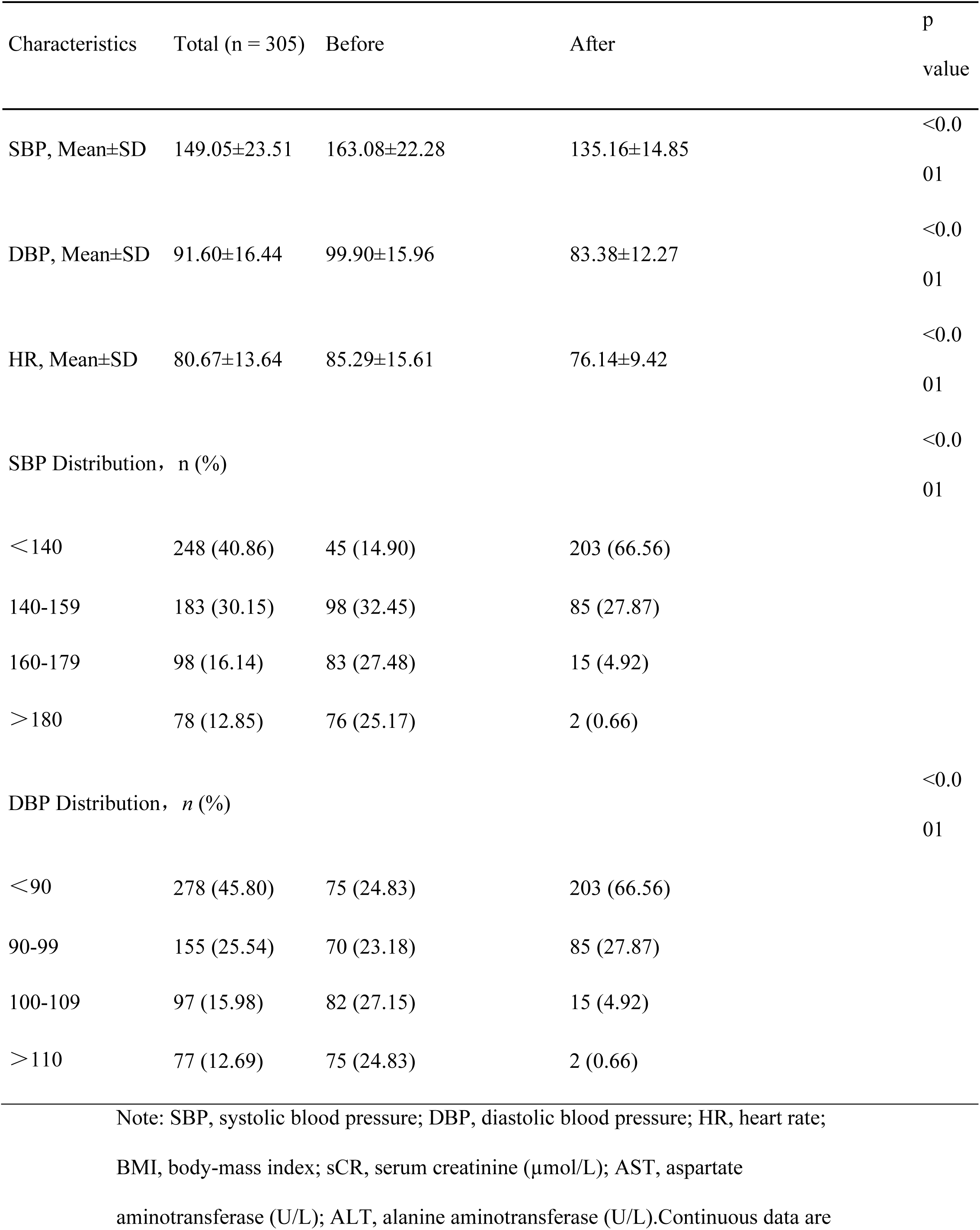
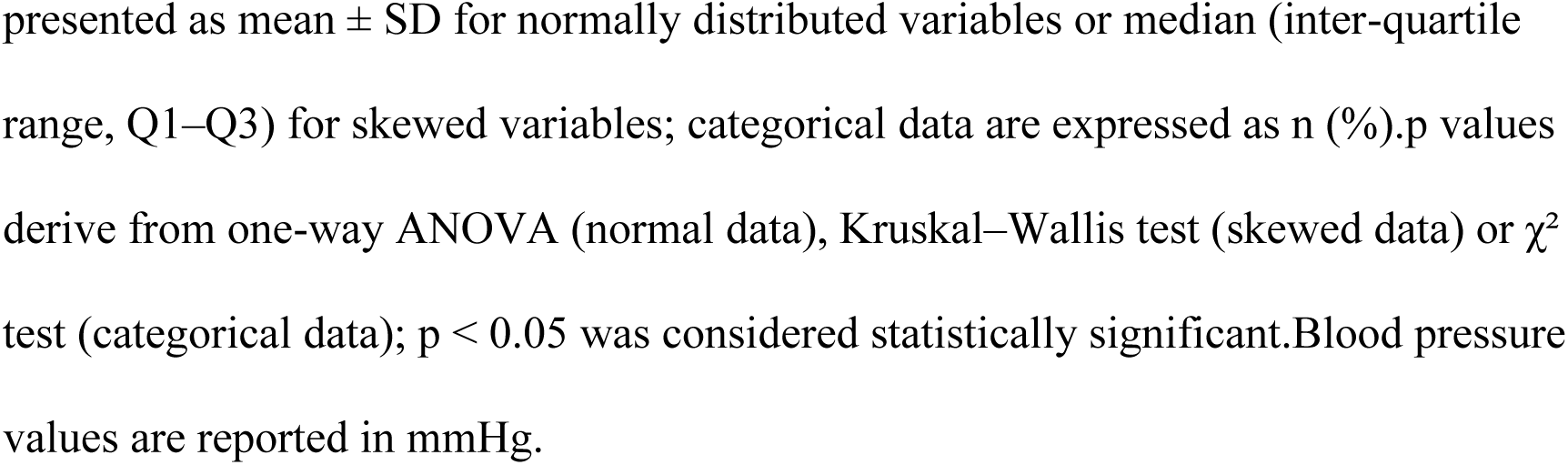
Comparison of Blood Pressure Before and After Adjusting Dual CCB Antihypertensive Therapy.

Among patients not previously treated with antihypertensive medication, the dual CCB regimen produced marked reductions in SBP (185.40 ± 19.40 vs. 138.82 ± 15.96 mmHg; p < 0.001), DBP (112.21 ± 14.84 vs. 86.31 ± 12.51 mmHg; p < 0.001), and HR (89.11 ± 15.67 vs. 76.21 ± 8.35 bpm; p < 0.001). Adding verapamil to amlodipine monotherapy further enhanced blood pressure reduction (p < 0.001) and modestly lowered HR (84.57 ± 15.48 vs. 79.35 ± 11.15 bpm; p = 0.047). Compared with regimens combining an angiotensin-converting enzyme inhibitor (ACEI), angiotensin receptor blocker (ARB), or angiotensin receptor– neprilysin inhibitor (ARNI) with a CCB, the dual CCB approach was significantly more effective in lowering SBP (158.77 ± 18.24 vs. 128.78 ± 14.42 mmHg; p < 0.001), DBP (95.80 ± 14.88 vs. 80.11 ± 11.43 mmHg; p < 0.001), and HR (85.21 ± 14.21 vs. 75.89 ± 7.69 bpm; p = 0.002). Similarly, compared with ACEI/CCB plus β-blocker therapy, dual CCB treatment provided superior SBP (152.19 ± 20.02 vs. 134.38 ± 12.45 mmHg; p = 0.001) and DBP (93.86 ± 12.54 vs. 83.19 ± 10.36 mmHg; p = 0.005) reduction, with equivalent HR control (75.40 ± 14.87 vs. 77.38 ± 10.90 bpm; p = 0.628).

Even among patients previously requiring three or more antihypertensive agents, the dual CCB combination remained effective, reducing SBP (154.90 ± 19.65 vs. 134.94 ± 14.48 mmHg; p < 0.001), DBP (92.25 ± 15.14 vs. 81.33 ± 12.85 mmHg; p < 0.001), and HR (83.10 ± 17.91 vs. 74.81 ± 9.45 bpm; p = 0.004). These findings underscore its potential role in resistant hypertension (Figure 3). Collectively, the evidence shows that dual CCB therapy is both safe and effective, achieving target blood pressure levels while meeting the requirements for medication washout (Figure 2).

**Figure 2.**
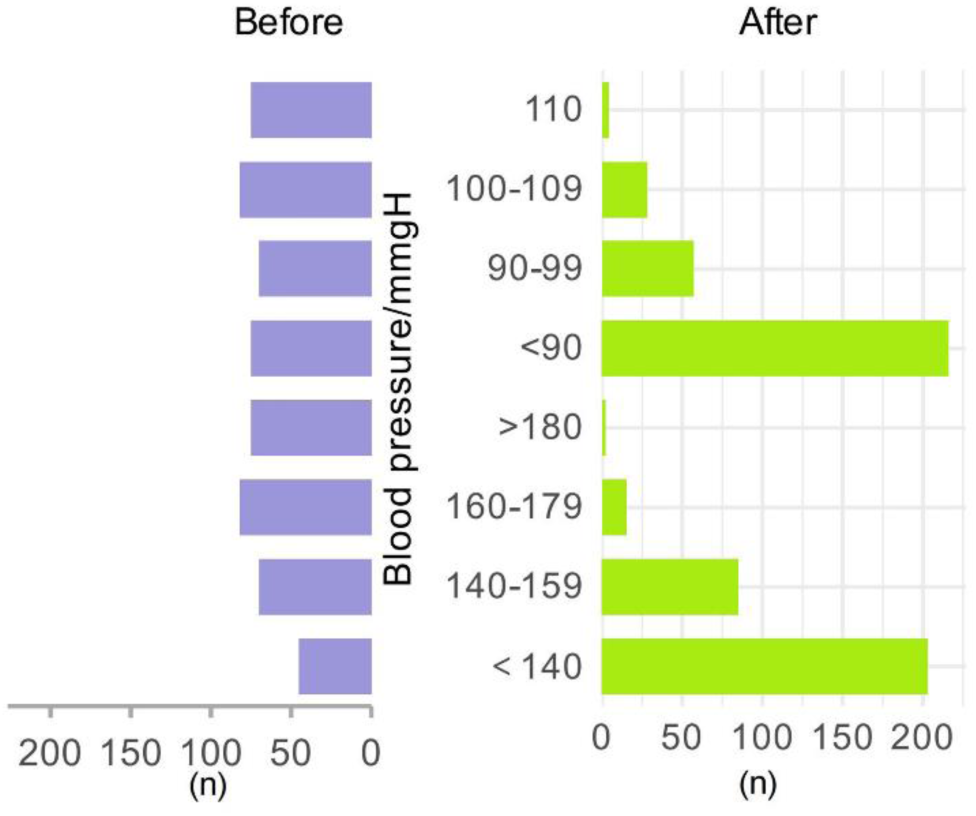
Blood Pressure Distribution Before and After Dual CCB Therapy.

**Figure 3.**
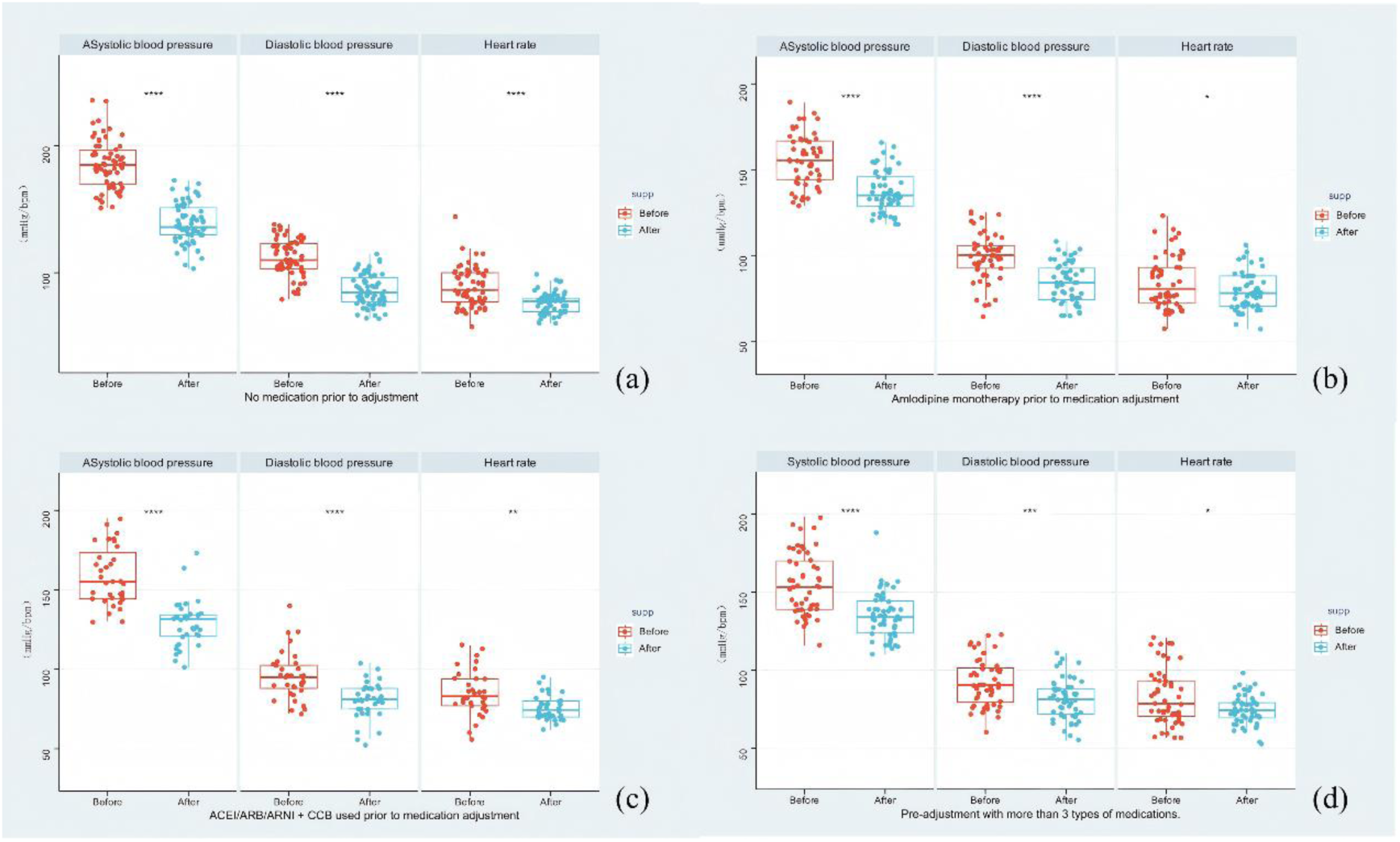
Antihypertensive efficacy of dual CCB combination therapy (amlodipine + verapamil).

### Adverse Reactions

During the washout phase, one patient developed intermittent second-degree type I atrioventricular (AV) block after receiving verapamil. This patient had no notable cardiac history and was not taking other medications known to affect cardiac conduction. Electrocardiography (ECG) confirmed the AV block after drug administration. Upon discontinuation of verapamil, the patient’s ECG returned to normal, heart rate stabilized, and no further symptoms occurred. Continuous cardiac monitoring detected no additional abnormalities, and the patient’s condition did not worsen.

## Discussion

The aldosterone-to-renin ratio (ARR) remains the most sensitive screening tool for primary aldosteronism (PA)[6].However, antihypertensive medications can interfere with its interpretation [7]. Abrupt withdrawal of these drugs without proper oversight may expose patients to cardiovascular risks, while a carefully designed washout protocol can balance safety with diagnostic accuracy. Non-dihydropyridine calcium channel blockers (CCBs) and α-blockers are generally believed to have minimal effects on ARR. Even so, adjusting treatment to include only these drugs is feasible for only about half of patients and may precipitate serious complications, such as hypertensive crises or atrial fibrillation[8]. These limitations highlight the need for a safer and more broadly applicable washout strategy.

Historically, amlodipine was excluded from washout protocols because of concerns that it might cause false-negative ARR results. This perspective has shifted. The 2020 guidelines from the Italian Society of Hypertension (SIIA)[9] indicate that its impact on ARR is minor, and while it may lower ARR values, the chance of producing false negatives is extremely low [10]. At the same time, there is growing consensus that all hypertensive patients should be screened for PA[6]. In China, amlodipine is widely prescribed as a first-line therapy under national hypertension guidelines. Compared with α-blockers, it has broader indications and is more widely used. This evolving view suggests that for patients with lower baseline blood pressure, the washout step could be minimized, simplifying the overall screening process.

The rationale for combining two CCBs is grounded in their complementary pharmacological properties. Both amlodipine and verapamil block L-type calcium channels, but they act differently [11, 12]. Amlodipine targets vascular smooth muscle, lowering resistance and blood pressure by dilating peripheral arteries. Verapamil, by contrast, acts in a “use-dependent” manner, becoming more effective at higher myocardial depolarization rates [13]. It reduces blood pressure mainly by slowing sinoatrial conduction and lowering ventricular rate, with less effect on vascular tone. As a result, the sympathetic activation and reflex tachycardia often triggered by amlodipine [14, 15] can be offset by verapamil’s negative chronotropic effect. In our study, patients receiving amlodipine alone experienced further blood pressure and heart rate reductions when verapamil was added, in line with these pharmacological mechanisms. Importantly, while amlodipine can induce peripheral edema in a dose-dependent manner [16], we observed no increase in edema after adding verapamil, supporting the view that their adverse effect profiles are distinct.

This study is one of the largest real-world analyses to examine dual CCB therapy in the setting of antihypertensive washout. Interestingly, patients without PA tended to have higher baseline blood pressure than those with PA. This may reflect selection bias, as patients with more severe hypertension are more often referred for screening, while some PA cases may remain in subclinical stages and evade detection.

Differences in prior medications may also have contributed. To reduce these confounders, we applied propensity score matching (PSM). Our results show that a dual CCB regimen of amlodipine plus verapamil provides safe and effective blood pressure and heart rate control during washout, comparable to the “dual CCB + α -blocker ” strategy. A possible explanation is that verapamil reduces plasma norepinephrine[17], producing effects that overlap with α-blockade. At the same time, the strong vasodilatory effect of two CCBs may mask any additional benefit from α-blockers. While both amlodipine and verapamil can theoretically suppress aldosterone production by blocking calcium channels involved in adrenal regulation[18–21], our findings did not demonstrate superior antihypertensive outcomes in PA patients. This may suggest that the anti-aldosterone effects of CCBs are insufficient to influence clinical results. Further studies are needed to clarify these mechanisms[22].

In hypertension management more broadly, international guidelines traditionally recommend dual therapy with renin–angiotensin system (RAS) inhibitors, such as ACE inhibitors or ARBs, combined with a CCB[23]. This strategy is favored for its organ-protective effects, neutral metabolic profile, and tolerability[24]. Our findings challenge this model by showing that a dual CCB regimen not only achieves superior blood pressure control compared with ACEI/ARB + CCB combinations but also reduces resting heart rate, with good safety. Notably, dual CCB therapy remained effective even in patients previously requiring three or more agents, suggesting a role in resistant hypertension. Despite this, current guidelines in China, the United States, and Europe do not recommend dual CCB therapy[23, 25]. Earlier studies labeled this approach as “immature” due to small sample sizes, lack of standardized dosing, and uncertainty about whether benefits reflected true synergy or dose effects [15]. As treatment strategies have shifted toward “low-dose combination” therapy, supported by trials such as QUARTET[26], it has become clear that combining agents with different mechanisms can maximize benefits without requiring high doses. Our results support reevaluating the role of dual CCB therapy within modern treatment strategies.

This study has several limitations. First, it was a single-center, retrospective analysis. Although the sample size was larger than previous reports, reliance on electronic records introduces potential selection and information bias. Multi-center studies would strengthen the findings. Second, follow-up was limited to the washout period; long-term cardiovascular and metabolic outcomes remain unknown. Third, because verapamil is a moderate CYP3A4 inhibitor, drug concentration monitoring for both verapamil and amlodipine would have been useful but was not performed. Finally, adverse events were recorded only through ward and outpatient notes. Without standardized prospective monitoring, events such as mild edema or constipation may have been underreported. Only one case of conduction block was identified, but routine electrocardiography, echocardiography, and edema assessments were not conducted, further limiting safety data. Future studies should address these gaps with more comprehensive monitoring.

Taken together, our findings suggest that the combination of amlodipine and verapamil is a practical and effective option for antihypertensive washout before PA screening. Based on our clinical experience, it can be applied broadly, offering a safe and efficient strategy for patients. With additional research and larger datasets, dual CCB therapy may emerge as an important component of hypertension management, particularly for resistant cases. Future work should focus on its long-term efficacy, safety, and potential synergies with other antihypertensive agents.

## Conclusion

The dual CCB regimen of amlodipine and verapamil demonstrates strong antihypertensive effects with good tolerability. It can be widely applied for drug washout before PA screening and may offer added value in resistant hypertension. Further multi-center studies are needed to confirm its long-term safety and efficacy and to explore how best to integrate this approach into combination therapy strategies.

## Data Availability

Data availability: The data that support the findings of this study are not publicly available due to privacy concerns. However, the data are available from the corresponding author upon reasonable request.

